# Predicting Diabetes in Canadian Adults Using Machine Learning

**DOI:** 10.1101/2024.02.03.24302302

**Authors:** Kayla Esser, Monica Duong, Khalil Kain, Son Tran, Aryan Sadeghi, Aziz Guergachi, Karim Keshavjee, Mohammad Noaeen, Zahra Shakeri

## Abstract

Rising diabetes rates have led to increased health-care costs and health complications. An estimated half of diabetes cases remain undiagnosed. Early and accurate diagnosis is crucial to mitigate disease progression and associated risks. This study addresses the challenge of predicting diabetes prevalence in Canadian adults by employing machine learning (ML) techniques to primary care data. We leveraged the Canadian Primary Care Sentinel Surveillance Network (CPCSSN), Canada’s premier multi-disease electronic medical record surveillance system, and developed and tuned seven ML classification models to predict the likelihood of diabetes. The models were tested and validated, focusing on clinical patient characteristics influential in predicting diabetes. We found XGBoost performed best out of all the models, with an AUC of 92%. The most important features contributing to model prediction were HbA1c, LDL, and hypertension medication. Our research aims to aid healthcare professionals in early diagnosis and to identify key characteristics for targeted interventions. This study contributes to an understanding of how ML can enhance public health planning and reduce healthcare system burdens.

## I. Introduction

Diabetes, a chronic metabolic disease characterized by hyperglycemia, is one of the largest global health emergencies of the 21st century[1], [2]. Diabetes rates continue to rise worldwide, as do the number of people experiencing acute and chronic complications. High blood glucose is estimated to be the third highest risk factor for premature mortality globally [3]. Canada has also seen rising rates of diabetes. As of 2022, 8.9% of the population had been diagnosed with diabetes, and prevalence has increased by an average of 3.3% per year[4], [5]. Diabetes is the leading cause of blindness, non-traumatic amputation, and end-stage renal disease in Canadian adults [3]. The cost of diabetes in Canada was estimated at $15.36 billion in 2022 [6]. However, it is also estimated that half of diabetes cases go undiagnosed, meaning the true population burden is higher than reported [7]. Early and accurate diagnosis and treatment of diabetes are imperative to prevent further disease progression and complications such as diabetic retinopathy, cardiovascular events and mortality [1].

Machine learning (ML) involves the application of computer algorithms to create a model of sample data to make predictions or decisions [8]. These models can be improved through testing, parameter tuning and validation. Deep learning is a subcategory of ML in which a neural network uses a hierarchical architecture to adapt to features of the dataset and learn from the data to improve predictive abilities. ML models have been used in the prediction, classification, and management of diabetes [1], [8]–[17]. Models that can predict diabetes may be useful for aiding clinician diagnosis of diabetes, as well as highlighting which features may be meaningful to target for early intervention. Much of the literature on ML diabetes prediction has focused on the US and other populations, therefore a model developed and validated on Canadian patient data may be more informative for the Canadian context, given differences in the health care system [14].

This study uses cohort data from a Canada-wide multi-disease electronic medical record surveillance system to answer the following research questions (RQs):

**RQ1:** Can we predict diabetes prevalence accurately, by comparing the performance of seven ML models: logistic regression (LR), decision tree (DT), random forest (RF), XGBoost (XGB), support vector machine (SVM), combined Naive Bayes (CNB), and an artificial neural network (ANN)? **RQ2:** Which clinical patient characteristics are most influential in the best-performing ML model’s prediction of diabetes prevalence?

These research questions have significant implications for public health planning and interventions. Our goal is to aid physicians in accurate diagnosing of diabetes, which can lead to earlier treatment and intervention, and ultimately reduce disease burden and health care system costs. Additionally, a systematic review of predictive machine learning diabetes models found a large variety in both the number and type of features included in models, indicating a lack of consensus [1]. Our study may clarify the utility of clinical and biomarker data available in the health records of many patients.

## II. Methods

### A. Data collection and preparation

This study used data from the Canadian Primary Care Sentinel Surveillance Network (CPCSSN), Canada’s first multi-disease electronic medical record surveillance system [18]. The CPCSSN is a network that spans eight provinces and one territory and has over two million patients and 1500 primary care clinicians [18]. CPCSSN extracts de-identified electronic medical records from primary care practices in Canada and applies cleaning, coding, and standardization algorithms to transform the data for use in quality improvement, surveillance, and research [18]. Diabetes cases are defined as type 2 diabetes mellitus, controlled or uncontrolled, excluding gestational diabetes, chemically induced diabetes, neonatal diabetes, hyperglycemia, prediabetes, or similar states or conditions [19]. The dataset for this study was generated by CPCSSN and shared with the research team. This was done by pulling patients aged 18 and up with a blood pressure reading and joining records that were the closest in time for other measurements (e.g., ±1 year). Patients on insulin were removed. The final data set is a random subset of 10,000 observations from the original CPCSSN dataset who had a blood pressure reading at their primary care providers’ offices between 2004 and 2014.

Data exploration included examining the extent of missing data in each variable, the variables’ distribution and central tendencies, and observing continuous variables’ correlations for potential collinearity. Missing values were addressed through multiple imputations by chained equations (MICE) for variables with under 10% missing (this cutoff was chosen to minimize introduced bias) [20]. We performed data standardization. In the case of two collinear variables, we selected one based on the strength of the association with the outcome in the literature. The final predictor variables used in all models were age, sex, systolic blood pressure (sBP), body mass index (BMI), low-density lipoprotein (LDL), high-density lipoprotein (HDL), HbA1c, triglycerides (TG), depression, hypertension, osteoarthritis, chronic obstructive pulmonary disease (COPD), use of hypertension medications and use of corticosteroids. For patients with multiple appointments, only data from the last visit was included. The final dataset contained 8,602 observations.

### B. Development of predictive models

We first held out a random 15% of the data for validation. Then, we randomly split the remaining data into 70% training and 30% testing. We used the training data to train the seven models (LR, SVM, DT, RF, XGBoost, CNB and ANN). Subsequently, we used the testing data to evaluate how well the models perform on unseen data (via confusion matrices and utility functions for accuracy, precision, recall, F1 score, and area under the receiver operating characteristic (ROC) curve (AUC)). A 5-fold cross-validation with grid-search was used to tune model hyperparameters to obtain the set of optimal hyperparameters that yielded the highest AUC. We tested multiple ANN models with varying configurations (number of hidden layers and type of regularization) to compare performance and reported the results for the ANN with the highest accuracy. We selected the best model based on an overall assessment of performance metrics and validated it on the held-out dataset to approximate external validation. We also conducted a feature importance analysis of the best model using SHAP (SHapley Additive exPlanations).

To ensure replicability and facilitate further research, the source code of all the presented machine learning models is available on GitHub^1^.

## III. Results

A total of 4,412 (48%) diabetic and 4,460 (52%) non-diabetic patients were included in this dataset for a total of n = 8,872 unique adult patients. The patients in the diabetic dataset were 51% male, with a mean age of 65 years (standard deviation [SD] 12 years), and those in the non-diabetic dataset were 49% with a mean age of 61 (SD 14) (see Table I for participant demographics). The majority of diabetic individuals (83%) were prescribed hypertension medication, compared to nearly half (54%) in non-diabetic individuals. Patients with diabetes presented a higher average concentration of HbA1c (mg/dL) levels.

**TABLE I:** Clinical patient characteristics. Data are mean [SD] or counts (%). P-values generated from t-test.

| Variable | Diabetic | Non-diabetic | p-value |
| --- | --- | --- | --- |
| n (%) | 4142 (48) | 4460 (52) | – |
| Age at exam (years) | 65 [12] | 61 [14] | < 0.001 |
| Gender (males) | 2104 (51) | 1885 (42) | < 0.001 |
| sBP (mmHg) | 131.2 [16.8] | 129.0 [17.0] | < 0.001 |
| Body Mass Index | 31.6 [7.0] | 29.1 [6.5] | < 0.001 |
| LDL (mmol/L) | 2.2 [0.9] | 2.9 [0.9] | < 0.001 |
| HDL (mmol/L) | 1.2 [0.4] | 1.4 [0.4] | < 0.001 |
| HbA1c (mg/dL) | 6.8 [1.1] | 5.7 [0.4] | < 0.001 |
| Triglycerides (mmol/L) | 1.7 [1.1] | 1.4 [0.9] | < 0.001 |
| Depression | 828 (20) | 981 (22) | 0.007 |
| Hypertension | 2941 (71) | 2368 (53) | < 0.001 |
| Osteoarthritis | 1364 (33) | 1264 (28) | < 0.001 |
| COPD | 437 (11) | 387 (28) | 0.003 |
| Hypertension Med | 3422 (83) | 2429 (54) | < 0.001 |
| Corticosteroids | 1182 (29) | 1221 (27) | 0.231 |

Missing data: n (D:ND): sBP 4 (2:2), LDL 52 (33:19), HDL 68 (20:48), TG 51 (17:34).

The optimal set of hyperparameters for each model is summarized in Table II. From the seven predictive models tested, we deemed the tuned XGBoost classifier as the best-performing model based on its high AUC (92%) (Figure 1; Table III). The XGBoost model was then tested on the held-out validation set. This yielded the following performance metrics in non-diabetic and diabetic patients: 80% precision (ND), 87% recall (ND), F1 score of 83% (ND), 87% precision (D), 79% recall (D), F1 score 83% (D), 83% accuracy, AUC 90%, and a macro-average of 83%. SHAP analysis of the XGBoost model revealed the five most influential features were HbA1c, LDL, hypertension medication, HDL, and BMI (mean SHAP values 2.10, 0.34, 0.22, 0.16, and 0.14, respectively) (Figure 2).

**TABLE II:** Classifiers and their tuned hyperparameters.

| Model | Hyperparameters | Parameters | Selections |
| --- | --- | --- | --- |
| LR | C<br>penalty<br>solver | [0.01, 0.1, 1, 10]<br>['l1', 'l2']<br>['liblinear', 'saga'] | 1<br>'l1'<br>'liblinear' |
| DT | max_depth<br>min_samples_leaf | [5, 10, 15, 20]<br>[5, 10, 15, 20] | 5<br>10 |
| RF | n_estimators<br>max_depth<br>min_samples_leaf | [10, 50, 100, 200]<br>[5, 10, 15]<br>[15, 20, 25] | 200<br>15<br>15 |
| XGBoost | n_estimators<br>learning_rate<br>max_depth<br>gamma<br>reg_lambda<br>reg_alpha | [50, 100, 200]<br>[0.01, 0.1, 0.2]<br>[3, 5, 7]<br>[0, 0.1, 0.2]<br>[0, 1, 5]<br>[0, 1, 5] | 100<br>0.1<br>3<br>0.2<br>5<br>1 |
| SVM | kernel<br>C<br>gamma<br>class_weight | ['rbf']<br>[0.01, 0.1, 1, 10]<br>[0.01, 1, 10]<br>[None, 'balanced'] | 'rbf'<br>1<br>0.01<br>'balanced' |
| CNB | GNB: smoothing<br>BNB: alpha | np.logspace(0, -9, 10)<br>np.logspace(0, -9, 10) | 0.001<br>1 |
| ANN | layers<br>hidden_units<br>lambda<br>epochs<br>learning_rate<br>LR_scheduling<br>batch_size<br>regularization | [1, 2]<br>[64, 128, 256]<br>[0.001, 0.01, 0.1]<br>[500, 1000]<br>[0.0001, 0.001, 0.01, 0.1]<br>[0.9]<br>[32, 64, 128]<br>['dropout', 'l2'] | 2<br>128<br>0.01<br>500<br>0.001<br>0.9<br>64<br>'l2' |

**TABLE III:**
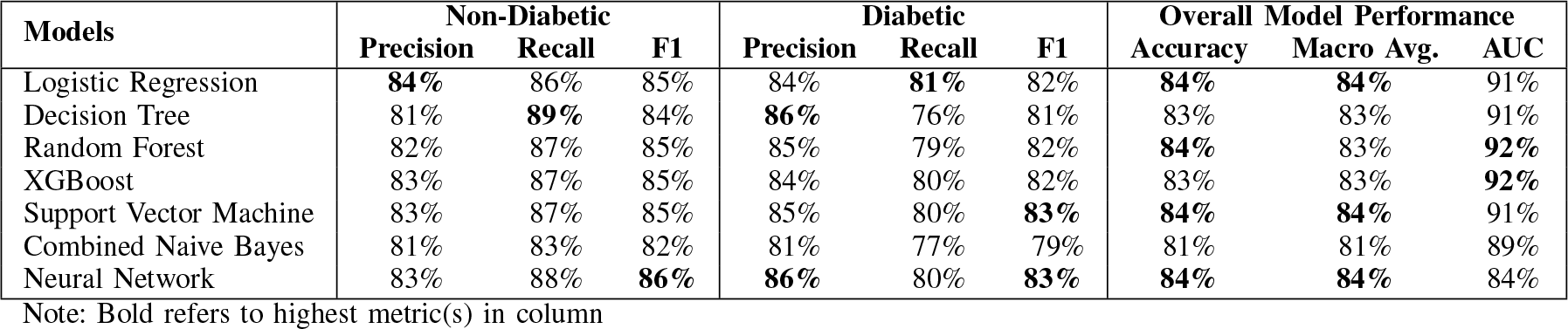
Comparison of performance metrics between LR, DT, RF, XGB, SVM, CNB and ANN algorithms in predicting diabetes prevalence in the test dataset.

| Models | Non-Diabetic |  |  | Diabetic |  |  | Overall Model Performance |  |  |
| --- | --- | --- | --- | --- | --- | --- | --- | --- | --- |
|  | Precision | Recall | F1 | Precision | Recall | F1 | Accuracy | Macro Avg. | AUC |
| Logistic Regression | **84%** | 86% | 85% | 84% | **81%** | 82% | **84%** | **84%** | 91% |
| Decision Tree | 81% | **89%** | 84% | **86%** | 76% | 81% | 83% | 83% | 91% |
| Random Forest | 82% | 87% | 85% | 85% | 79% | 82% | **84%** | 83% | **92%** |
| XGBoost | 83% | 87% | 85% | 84% | 80% | 82% | 83% | 83% | **92%** |
| Support Vector Machine | 83% | 87% | 85% | 85% | 80% | **83%** | **84%** | **84%** | 91% |
| Combined Naive Bayes | 81% | 83% | 82% | 81% | 77% | 79% | 81% | 81% | 89% |
| Neural Network | 83% | 88% | **86%** | **86%** | 80% | **83%** | **84%** | **84%** | 84% |

Note: Bold refers to highest metric(s) in column

**Fig. 1:**
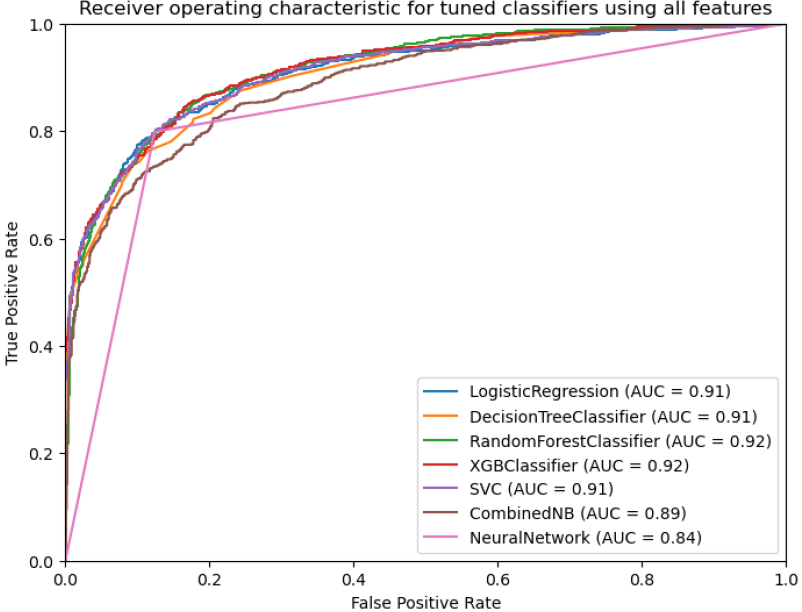
ROC curves for tuned classifiers and neural network models on the test dataset.

**Fig. 2:**
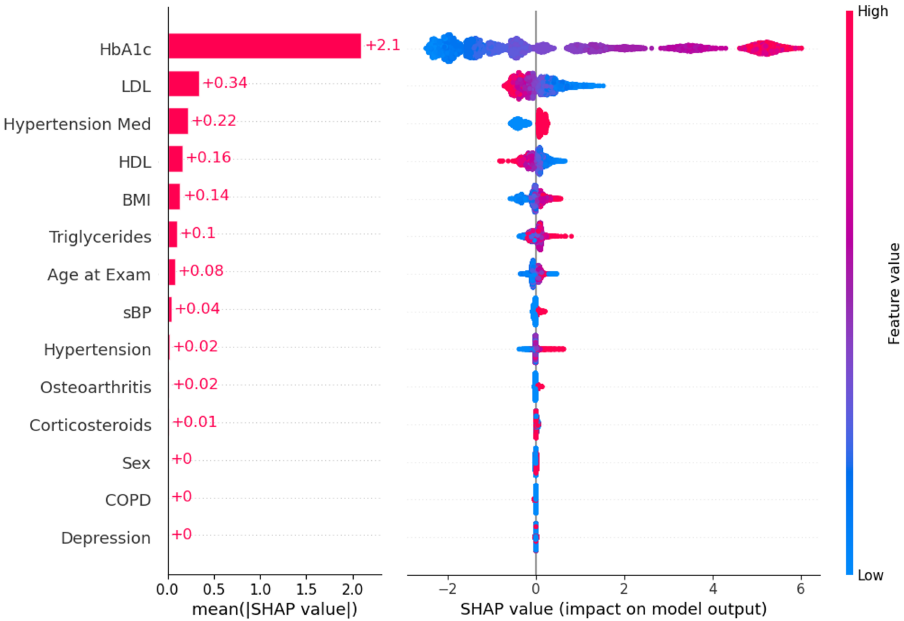
SHAP summary plot and mean values for predicting diabetes using the tuned XGBoost model.

## IV. Discussion

The main goal of this study was to determine the best model to predict diabetes prevalence by comparing the performance of seven ML models and identifying the most influential clinical patient characteristics in the model’s predictions. The use of seven ML model architectures allowed for a comprehensive comparison of methods to evaluate predictive accuracy and recall, while limiting overfitting through the use of cross-validation. We observed consistent predictive performance of the tuned XGBoost classifier across the training, testing, and validation datasets. Furthermore, visual inspection of the ROC curves suggests that the XGBoost classifier has a smoother shape. This indicates less instability in its predictions and thus less overfitting. We identified XGBoost as the best-performing model based on these characteristics. The high AUC indicated the model performs well at distinguishing positive and negative diabetes classes, and the validation results mean the model can generalize well to real-world clinical settings compared to the other classifiers. These findings align with existing literature on diabetes prediction using ML. One study compared SVM, K-Nearest Neighbours (KNN), NB, DT, LR, and XGBoost algorithms for diabetes prediction using physical examination data and found XGBoost had the highest accuracy (81%), similar to our model’s accuracy of 83% [21]. In another study by Li et al. (2020), the authors also used CPCSSN data to compare the performance of LR, Gradient Boosting Method (GBM), DT, and RF models in predicting diabetes, using parameters age, BMI, TG, FBS, sBP, HDL, and LDL [14]. They found GBM and LR outperformed RF and DT, with 85% and 84% AUC and 72% and 73% sensitivity, respectively [14]. Our XGBoost model has higher AUC and sensitivity (recall) scores than the other study’s GBM model, as XGBoost is a more regularized form of GBM which contributes to improved model generalization capabilities [22]. Our models also included additional clinical features, which could contribute to improved performance.

The SHAP analysis the most important features for prediction. For example, a mean SHAP value of 2.10 for HbA1c implies that this feature contributes positively to the model’s prediction across multiple instances (Figure 2). This may have useful clinical applications as these features could be prioritized for clinical monitoring of patients at risk of developing diabetes. When validating the model on unseen data, the AUC (91%) and accuracy (83%) remained high. This suggests the model generalizes fairly well, indicating that it has learned patterns that are applicable beyond the training data.

Our XGBoost model could potentially serve as a valuable tool for physicians in diagnosing or identifying the risk of diabetes in Canadian patients. The current standard procedures for identifying prediabetes typically rely on an FBS range of 6.1-6.9 [23]. However, HbA1c might offer a more comprehensive assessment. Unlike FBS, HbA1c reflects an average of blood sugar levels over the past 2-3 months, providing a more holistic view of glycemic control [24]. By incorporating HbA1c along with other traditionally significant features in diabetes onset, our model aims to enhance diagnostic accuracy. This refinement could potentially lead to more precise identification and management of individuals at risk of developing diabetes. Additionally, the results of our study are likely to be generalizable given the large, nationally representative sample.

Several limitations of this work should be considered. Firstly, this study was conducted retrospectively, therefore causation cannot be inferred based on the associations found by our predictive models. Secondly, data for this analysis was only captured until 2014, therefore it may not be reflective of current diabetes prevalence or risk factors. Thirdly, our dataset did not contain a race or ethnicity variable, which limited our ability to explore the impact of race or ethnicity on diabetes prevalence. Future work should use a timely database and investigate differential risks for various sex, ethnicity, and socioeconomic groups, as marginalized populations have been demonstrated to experience disproportionately higher diabetes risk [25].

## V. Conclusion

This study used Canada-wide surveillance data to evaluate and compare seven machine learning models to predict diabetes prevalence and identify specific clinical characteristics most important in this prediction. Our analysis identified the XGBoost model as the top-performing model and HbA1c, LDL, hypertension medication, HDL, and BMI as the top five most influential features. This may be disseminated into clinical settings to assist with diagnosis and risk identification, which can inform patient care and resource allocation. For example, interventions for preventing diabetes may consider targeting risk factors through diet, exercise or medication. Earlier diagnosis of diabetes can reduce disease burden and overall health system costs.

## Data Availability

All data used are available online at https://cpcssn.ca/ website upon reasonable request.

## Footnotes

1 https://github.com/andysontran/2024_IEEE_EMBC_Diabetes-I

